# Co-Identifying Policy-Relevant Modelling Questions: A Case Study of the Human Papillomavirus (HPV) Vaccine Introduction in Mozambique

**DOI:** 10.1101/2025.08.20.25334054

**Authors:** Paula Christen, Agostinho Viana Lima, Muanacha Mintade, Neusa Torres, Timothy Hallett, Allison Portnoy, Lesong Conteh

**Affiliations:** Medical Research Council Centre for Global Infectious Disease Analysis, Faculty of Medicine, Imperial College London, London, UK.; Manhiça Health Research Centre, Manhiça, Mozambique; Harvard Center for Health Decision Science, Harvard University, Boston, MA, USA; Department of Health Policy, London School of Economics, London, UK; LSE Health, London School of Economics, London, UK; Department of Global Health, Boston University School of Public Health, Boston, MA, USA

## Abstract

Mathematical models hold the potential to generate valuable evidence for shaping vaccination policies. However, maximizing their impact requires a deeper understanding of how modelling efforts can be aligned with the real-world priorities of policymakers and health officials. This study explores how structured engagement with stakeholders can help co-identify decision-relevant questions that are amenable to quantitative modelling. The focus is the human papillomavirus (HPV) vaccination programme in Mozambique.

We conducted semi-structured interviews with stakeholders involved in the HPV vaccine programme to identify key knowledge gaps in their decision-making context, i.e., practice. These were translated into research questions that informed the application of a mathematical model. An evidence brief was developed to synthesize and contextualize findings, and follow-up interviews were conducted to reflect on the utility of the evidence. Qualitative data were analysed inductively to identify emergent themes.

Stakeholders identified four priority questions: optimal vaccine delivery strategy, distributional impact, vaccine economics, and comparison with other prevention methods. They emphasized the value of tailored evidence—particularly at the provincial level—for informing financial planning, resource allocation, and advocacy. The approach facilitated collaboration between researchers and stakeholders, helped uncover previously untapped data sources, and improved the policy relevance of the modelling outputs.

This study demonstrates how co-identifying modelling questions with decision-makers can help ensure that evidence generated through mathematical models is context-specific, and policy-relevant. This type of engagement enabled clearer alignment between model development and decision-making needs—offering lessons for future applications of modelling in public health policy.

## 2. Introduction

Cervical cancer, a vaccine-preventable disease caused by the human papillomavirus (HPV), disproportionately affects women in East Africa, accounting for 10.7% of global mortality.^1^ While three prophylactic HPV vaccines targeting various HPV genotypes are available, with promising new candidates in development^2^, their introduction and implementation have been varied across countries across sub-Saharan Africa.^3^ Almost two-thirds of countries in the region have introduced the vaccine to their national immunization programme.^4^ Others remain in various stages of preparation or decision-making, despite recent commitments to expand vaccine access.^5^

From funding for research on a new vaccine to scaling it nationally, regionally, or globally, various types of evidence inform decisions for vaccine introduction at multiple levels.^6–9^ The relevance and appropriateness of different types of evidence for vaccination-programme-related decisions vary over time and by stakeholder.^10^ One type of evidence that can inform public health policy options can be developed with mathematical models. Mathematical models can be used to efficiently test experimental hypotheses that would otherwise be expensive, difficult or unethical to explore.^11^ As such, they have the potential to provide insights into policy questions^12^, assist in addressing complex policy issues, guide decision-making regarding resource allocation, and advocate for resources.^13–15^

Global burden of disease and vaccine impact estimates, in particular, hold a significant advocacy function and are often used by international organizations to garner financial resources.^15^ However, these global models have been criticized for their limited applicability to specific national and sub-national contexts, where policy and implementation decisions are made.^15^ Stakeholders at these levels often require more detailed and tailored models that can address their unique challenges and priorities.

The knowledge translation literature suggests that actively embedding evidence consumers (e.g., policymakers and public health officials) throughout the evidence development process can significantly enhance the relevance and applicability of evidence generated.^16–18^ Yet, the current landscape is marked by ad hoc and often haphazard approaches to the engagement of infectious disease modellers with evidence consumers. This has resulted in missed opportunities to learn about how engagement between mathematical modellers and evidence consumers materializes, who to involve in the process, as well as highlighted a need for a more systematic and intentional approach to stakeholder engagement.^19–22^

Stakeholder engagement initiatives, aiming to enhance models for use in health policy, operate with a dual focus: generating ‘better science’ and facilitating ‘better decisions’.^23^ By incorporating evidence consumers’ input, mathematical models can be tailored to address local concerns and challenges, increasing their utility and relevance.^24^ Engagement in the modelling process can ensure that ‘needs’ and ‘preferences’ of evidence consumers embedded in local communities are represented ^25^ and help parameterize models with realistic field data.^26^ As models are attuned to local contexts, their quality, outputs, salience and relevance can improve.^23^

Models can function as ‘tools’ to help build a shared understanding or ‘common “mental map” of the health problem’^27^, serving as a ‘rallying point’ around which stakeholders can gather.^28,29^ This shared understanding can, for example, be used by health planners as a catalyst for convening others in thinking about their strategic directions and policy priorities, ultimately aligning actions and fostering more informed decision-making.^30^ Engagement can also foster ownership and trust, enhancing the mutual learning, transparency, and understanding of research processes.^19^

This paper explores how mathematical modelling can be used to inform specific policy and implementation questions at national and sub-national levels, moving beyond global impact estimates. The study investigates an approach to evidence development in which researchers worked closely with policymakers and implementers to define relevant questions for modelling, interpret findings, and assess their utility for decision-making. The aims of this research are to: (1) identify and jointly frame research questions with stakeholders that reflect the evidence needs of the HPV vaccination programme in Mozambique; (2) develop a mathematical model addressing these questions using data from sources identified through stakeholder engagement and literature review; and (3) contextualize and interpret model findings with those responsible for programme implementation and national scale-up. This manuscript focuses on aims (1) and (3), while the modelling methods and results (aim 2) are reported elsewhere.^31^

## 3. Methods

### 3.1. Study setting

This study was conducted in Mozambique, a southern African country, classified as a low-income country by the World Bank. The country has a population of almost 34 million (in 2023).^32^ Geographically, Mozambique stretches along the southeastern coast of Africa and is divided into 10 provinces.

In Mozambique, cervical cancer is one of the leading causes of cancer death among women. In 2020, Mozambique had one of the highest estimated age-standardized cervical cancer incidence rates globally, 50.2 per 100,000.^33^ This incidence rate is 3.7 times higher than the estimated global average of 13.3 per 100,000 and 1.8 times higher than the estimate for the Eastern African region of 20.6 per 100,000.^1^

Access to cervical cancer screening and treatment in Mozambique is very limited. The cervical cancer screening programme is based on Visual Inspection with Acetic Acid and cryotherapy in selected health facilities since 2009.^34,35^ Radiotherapy has been provided since 2019 at two facilities in Mozambique (the Maputo Central Hospital and the Central Private Hospital), both located in Maputo.^36^ Before radiotherapy services were available in the country, less than 1% of patients received optimal guideline-adherent therapy and less than 3% received therapy with major deviations from guidelines.^37^ Case-fatality rates are also high due to a lack of timely, safe, and affordable surgical care.^38^ Cancer treatment is not covered by Mozambique’s health coverage benefit package^39^ and therefore poses a significant financial burden on the individual.

The introduction of the HPV vaccination in Mozambique was a protracted process (Figure 1). Political influence, particularly from national champions such as the First Lady as well as global organizations like Gavi, the Vaccine Alliance (Gavi), and the World Health Organization (WHO), played a pivotal role in prioritizing and introducing the vaccine. A successful demonstration programme in three provinces (2012–2016) further solidified the decision for nationwide introduction.^40^ However, despite plans for a 2019 launch, natural disasters, the global HPV vaccine supply shortage, and the COVID-19 pandemic caused significant delays. The vaccine (GARDASIL®, Merck & Co. Inc.) was finally introduced in 2021, initially targeting 9-year-old girls, with subsequent catch-up campaigns planned to reach older cohorts.^41^

**Figure 1.**
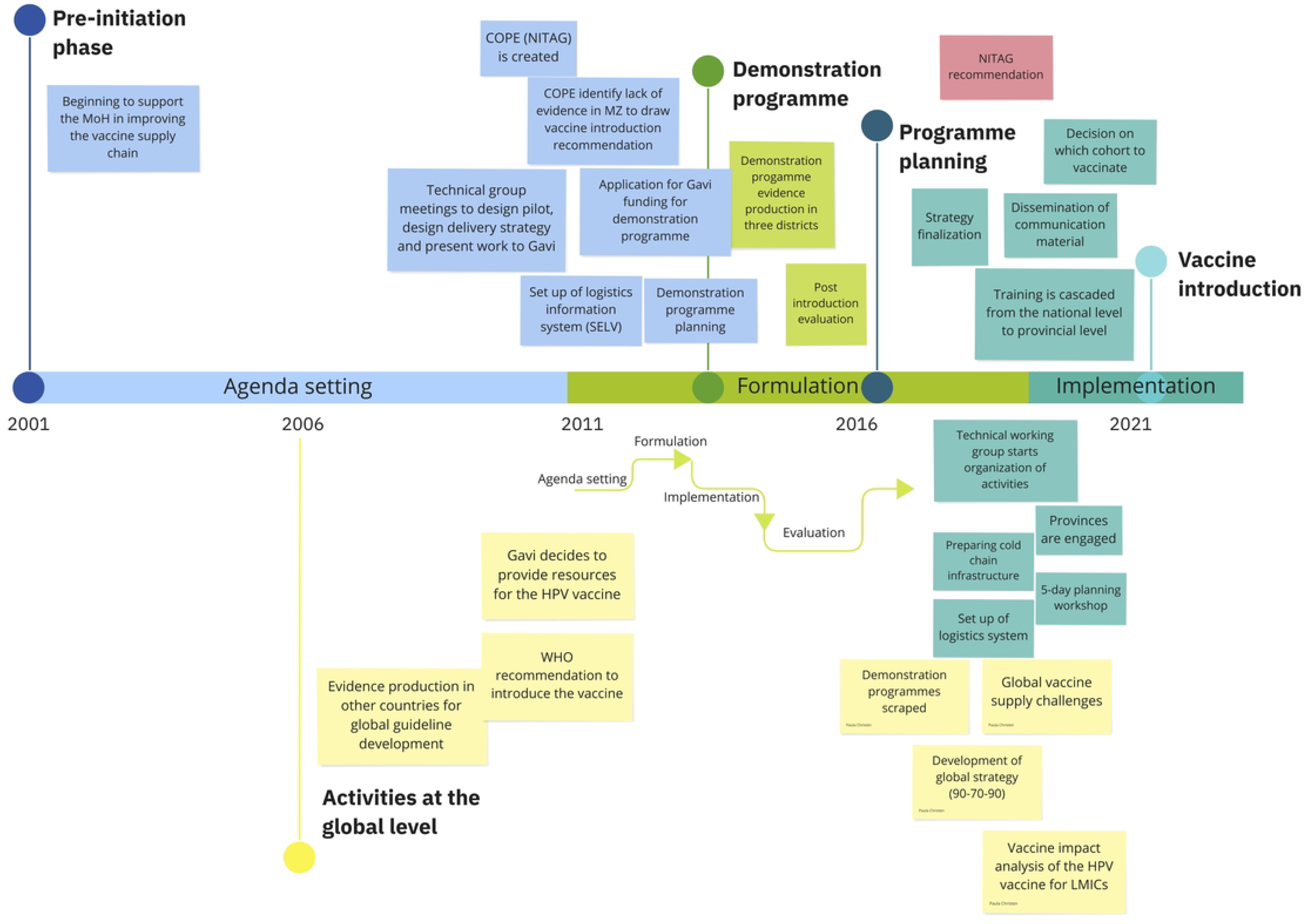
Chronology of HPV vaccination programme introduction in Mozambique. Timeline of key events related to the introduction of the HPV vaccine in Mozambique, categorized by stages of the policy cycle: pre-initiation, agenda setting, formulation, and implementation. Abbreviations: NITAG, National Immunization Technical Advisory Group; WHO, World Health Organization; HPV, human papillomavirus; NGOs, non-governmental organizations.

### 3.2. Study design

This study follows a mixed-methods approach (Figure 2). The findings from the qualitative components (Parts 1, 2 and 4) are presented here, while the evidence development (Part 3) is detailed elsewhere^31^ to maintain a focus on process rather than empirical modelling results.

**Figure 2.**
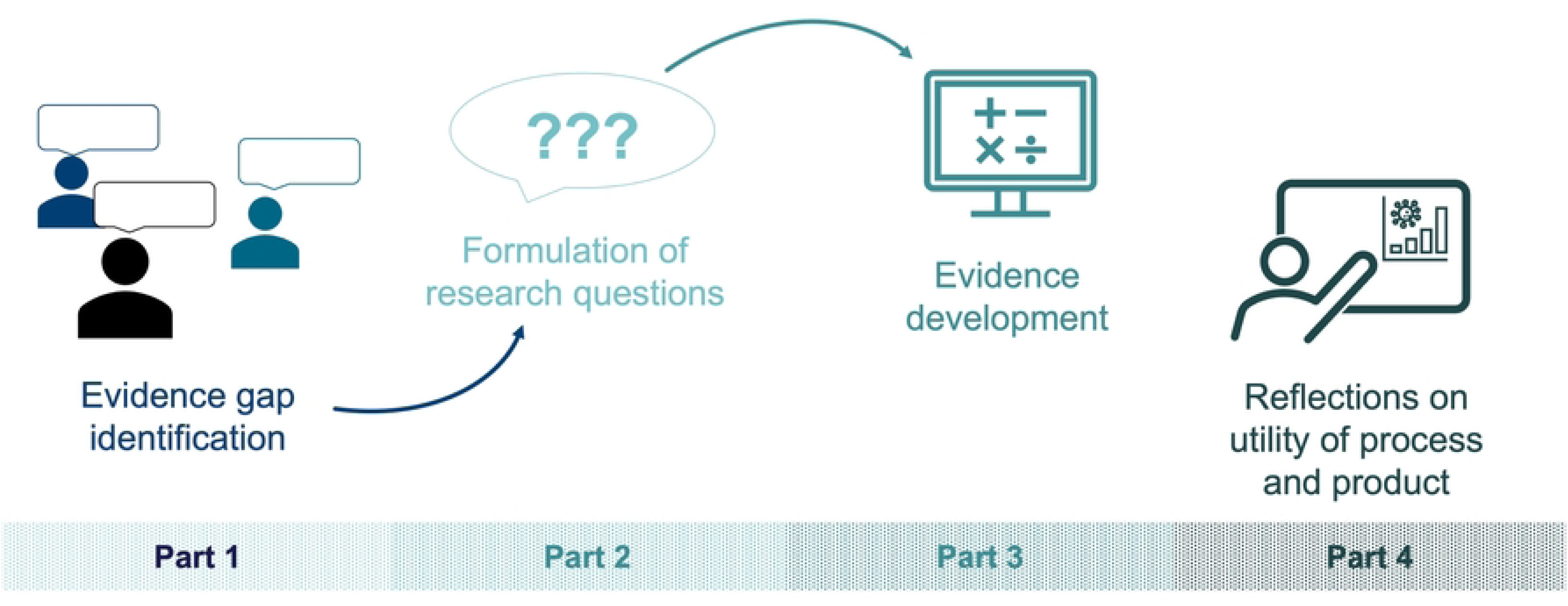
Overview of the methodology.

The co-creation process consisted of four parts: (1) identifying evidence gaps through stakeholder interviews, (2) formulating research questions based on priority themes, (3) developing evidence through modelling and analysis, and (4) reflecting on the utility of both the process and the resulting evidence products.

The study design was informed by prior research demonstrating that interviews with policy stakeholders are effective for identifying knowledge gaps and aligning research agendas with policy issues. Nyanchoka et al. ^42^ and others^43^ highlight the value of early engagement to ensure that evidence responds to the priorities of decision-makers. Crucially, literature across health and implementation science emphasizes that such engagement should be iterative, allowing researchers to refine and validate findings through repeated interactions.^44^ This approach—used in research-prioritization efforts^45^ and stakeholder-engaged programmes ^44^—has been shown to enhance the relevance, quality, and policy salience of research. Guided by these insights, we incorporated both initial and follow-up interviews to support the iterative identification and refinement of stakeholder-defined evidence needs.^20,44^

#### Part 1

Semi-structured interviews with open-ended questions were conducted to enable stakeholders to reflect on and describe aspects of the HPV vaccination programme in Mozambique that they wanted to know more about. The interview guide was co-developed and piloted with Mozambique-based research assistants, AL and MM (Supplement 1). During the interviews, mathematical models as a method to develop evidence were not mentioned by interviewers to avoid bias. Instead, the interview inquired about any form of evidence that could help stakeholders in the future.

Study participants were purposefully sampled to ensure we had representative views of those who had been engaged in the HPV vaccination programme planning by providing (1) financial investment or (2) technical support and/or (3) held formal decision-making power. They were identified through suggestions from Mozambican colleagues in the research team, an initial stakeholder mapping, and snowball sampling. Participants who met the inclusion criteria were associated with at least one of three broad categories: agenda setting, formulation and implementation (Figure 1). Study participants were categorized by the level at which their organization or office operated in the context of the HPV vaccination programme in Mozambique: headquarter (HQ), regional (R), national (N), provincial (P), and their type of organization: international organization (IO), non-governmental organization (NGO), Ministry of Health (MOH), other Ministry (M).

Interviews were conducted in English or Portuguese, depending on the interviewee’s preference, in person or via the telecommunication software Microsoft Teams (due to travel restrictions related to the COVID-19 pandemic) between 10 January 2022 and 30 May 2023. First round interviews were held between January 2022 and April 2022. Prior to interviews, study participants received a participant information sheet and were asked to sign an informed consent form. Interviews lasted between 25 and 120 minutes and were audiotaped. From the audio recordings, verbatim transcripts were produced. Interviews that were conducted in Portuguese were translated into English by MM. A sub-sample of interviews were then back translated to Portuguese as a quality assurance step. Study participants were pseudonymized to protect their identity; their type of affiliation and institution were their defining characteristics. This was done to enable people to speak more candidly. Transcripts were shared with study participants after the interview for them to clarify information or correct any transcription mistakes that arose due to low audio quality. Data collection via the first round of interviews was terminated once perspectives from individuals from all key organisations engaged in the HPV vaccination programme were probed.

#### Part 2

Building on the themes identified in Part 1, research questions were formulated by PC, MM and AA to reflect the evidence needs raised by participants and to identify areas that could be addressed through mathematical modelling. These questions were developed by the research team based on the gaps articulated during interviews, aiming to capture the intent and substance of participants’ concerns. With respect to the research questions elicited, tools or mathematical modelling approaches that had been applied in other countries or public health policy contexts were identified in the peer-reviewed literature by PC.

A subset of research questions was chosen. These questions were chosen considering the methods that can be used to address them, i.e., for the purpose of this research, mathematical models, and not based on their perceived priority (e.g., frequency across interviews). The questions were addressed by extending an existing HPV population-based epidemiologic model.^46^ The research group, which had developed this model, was engaged to explore the extension of the model to the national context of Mozambique. The model was parameterized with national data, where available, to estimate vaccine impact in the form of cervical cancer cases and deaths averted. The methodology to our extension of the modelling framework and findings have been published separately. Research questions and modelling results were presented in an evidence brief, a document that synthesizes and contextualizes scientific evidence with the aim to engage policymakers and present policy options.

#### Part 4

The evidence brief was shared with a subset of decision-makers involved in the vaccination programme planning and implementation originally interviewed. This subset was defined by those who raised knowledge gaps underlying the identified research questions. This subset of individuals was chosen to probe whether the findings adequately addressed these gaps.

Follow-up, semi-structured interviews were conducted to probe the policy relevance of research questions and investigate the utility of evidence in the form of vaccine impact estimates as presented in the evidence brief. Study participants from the first round of interviews were invited for these follow-up interviews, which took place in person and online in April and May 2023, a year after their original interview, if they still had the potential to influence decisions around the national HPV vaccination programme in Mozambique. By receiving the evidence brief prior to interviews, study participants could familiarize themselves with the content. During the interview, the evidence brief was explained in detail. Second round interviews were processed (transcribed, translated, and pseudonymized) in the same way as first round interviews. The data was coded inductively, through which emerging themes on feedback to the research questions, the evidence, and how the evidence could be used were identified.

Storage, organization, searching, and coding of transcripts from both interview rounds were performed using NVivo 12 Pro.^47^

Ethical approval was obtained from Imperial College London (ICREC reference: 21IC7033) and the Institutional Review Board of the Manhića Health Research Centre (CISBS-CISM/076/2021). Approval from the Bioethics Committee in Mozambique was not required for this study.

## 4. Results

Of the 50 individuals that were contacted for first round interviews, 28 (56%) participated (Supplement 2, Table 1). At the time of involvement, study participants were employees at organizations that had been engaged in the decision to introduce the HPV vaccine in Mozambique, the programme planning, and/or implementation. In addition, four study participants had been members of the Mozambican national immunization technical advisory group (NITAG), locally known as *Comitê de Peritos para Imunização (CoPI)*.

**Table 1.** Policymakers’ questions about the HPV vaccination programme in Mozambique that can be addressed using mathematical models. Abbreviations: M: Ministry; NGO: Non-governmental organization; IO: International organization; HQ: Global; N: National; P: Provincial

| Part 1 | Part 2 |  |
| --- | --- | --- |
| Interview extract | Knowledge gap translated to research question | Possible outcome measure(s) |
| **1. Vaccine delivery strategy and comparison to other cervical cancer prevention methods** |  |  |
| 1.1 N, IO: “Also the issue of sex. Because there was a lot of discussion about whether we could introduce to boys or give it only to girls, because boys can also transmit the disease and may have the disease and can transmit it to others it is true that they will not have cervical cancer because they do not have a cervix, but they can transmit the disease.” P28<br>P, M: “We are now vaccinating nine-year-old children. But we want to understand why it’s only nine years old and why just the girls and not the men. Are men not a source of HPV transmission?” P21 | 1.1 What would be the impact on population health if boys were also vaccinated? | Population-level burden of disease |
| 1.2 N, IO: “For me it is more in relation to the broad target groups in terms of age group, because we only covered 9-year-old girls, but we also wanted to cover more. [...] So in our context of sexual intercourse in some parts of the country it starts very early, we have premature marriages, so how do we align that with what we want.” P13 | 1.2 What impact does sexual debut prior to vaccination have on the population health benefit (e.g., if older age groups are included in the target group)? How should the target group be defined with respect to sexual intercourse? | Optimal target group that maximizes the population level impact in terms of burden of disease |
| 1.3 N, NGO: “Practically we’re not getting vaccines for the age group that we had initially intended. We’re just getting a single cohort. We’re going to do nine-year-old girls. So these are the things that we need to think about. These are the things that we need, structuring it like you would. And I think this is where the technical assistance needs to bring in these kinds of good practices into the ministry.” P13 | 1.3 What implications does the change in target cohorts from a multi-age-cohort strategy to a single-age-cohort strategy have? |  |
| 1.4 N, NGO: “It would also be important to have vaccination data and what is the level of immunization done so far, the question related to safe sexual practices, what is the tendency for condom use among adolescents and young people to also have this information about what the probability would be in a scenario where low vaccination and low rate of condom use, so what would that mean from the point of view of the incidence rate and prevalence of cervical cancer in these communities when trying to cross-check that data and see how it can work.” P14 | 1.4 How does the vaccine introduction relate to/interact with/compare with other public health interventions and safe sex practices (e.g., condom use)? | Comparison of cervical cancer elimination strategies and combinations of these |
| **2. Distributional impact of the vaccination programme** |  |  |
| 2.1 HQ, IO: “Then, going through with the programmatic people is what the potential delivery modes are and identify where girls at high risk are in the country. Is it in urban settings, in slum areas, so the issue is always to identify the high risk, and these are often in difficult-to-reach communities, literally in terms of geographic, but also just you can’t identify them because they are in the informal sector or yeah. And that is more of a problem compared to high income countries, right due to roll out of the vaccine.” P2<br>HQ, IO: “Hard-to-reach populations. We seem to be very bad at. We sometimes call them out of schoolgirls in southern Africa, for example. The programme is really school based, and it’s not very clear whether a very active attempt is made | 2.1 How do inequalities in access to education and infrastructure reflect in vaccination programme impact? | Distributional impact of the vaccination programme across population sub-groups |

| Part 1 |  | Part 2 |  |
| --- | --- | --- | --- |
| Interview extract |  | Knowledge gap translated to research question | Possible outcome measure(s) |
| to deliver out of school. So, you know, some girls, including those in private schools, may just miss out on the vaccine because they're not part of the public programme. That is quite well, you could call it unacceptable, but that's a shortcoming. So I think, who we are reaching remains a black, you know, a grey zone. Nomadic populations in some parts of Africa, you know, so research is needed in that direction." P4<br>N, IO: "The issue of access to rural-urban vaccines I still cannot see this in the data I access. if we were seeing that the coverage is only in urban areas and the rural areas is not having anything, we have to do to change the strategy. So I can't understand whether it's urban or rural." P13<br>HQ, IO: "So I think some kind of modelling around looking at socioeconomic inequity indicators against HPV coverage." P6 |  |  |  |
| **3. Vaccine economics** |  |  |  |
| 3.1 N, R: "We do not know yet what the cost effectiveness of this vaccine is here in Mozambique. So, doing this analysis now will allow us to say whether the vaccine that was introduced will be cost- effective during this implementation period and in addition we will not only analyze Gardasil, the one that was introduced, we will also analyze the other two vaccines for Cervarix and Gardasil 9. So this will allow us to know in the medium to long term if we make the decision to change vaccines and how to adopt it. Not because we are going to propose to change this vaccine that has already been introduced, but perhaps in the medium and long term the other vaccines give us more advantage or we have reached the conclusion, making sure that the vaccine that was introduced last year is more cost-effective compared to the others." P8 |  | 3.1 What is the cost effectiveness of the different vaccine candidates? | Cost-effectiveness ratio |
| 3.2 HQ, IO: "And then particularly for costing is whether countries have access to Gavi prices or not. That often also makes a huge difference when it comes to economic impact for the product itself." P2<br>N, M: "There is the aspect of the price of vaccines for us as managers and so we are taking into account that until now almost all the vaccines that we are using in our system are financed by partners and there always comes that fear of: what will happen if one day the partners do not want to pay for these vaccines." P10 |  | 3.2 What costs does the vaccination programme imply following the transition from Gavi funding? Or What financial resources/budgets are required for the HPV vaccination programme over time? | Vaccination programme costs |
| 3.3 N, IO: "If you reduce the number of women with cancer, you are also reducing the economic burden and investment in the area of treatment, so this is important to demonstrate." P13 |  | 3.3 What is the wider economic benefit of introducing the HPV vaccine? | Resource savings; financial risk protection provided |
| 3.4 N, IO: "I think that in the future, when there are projects of this nature, it is necessary that we can not only secure funds for the purchase of equipment, but we also have to secure funds for customs clearance, because if we do not deliver, if we sometimes look at those that are state budget funds, the Ministry of Health use it to clear it if it's not available so it takes time. Then it is also necessary in the budgets to make planning, to make forecasts in order to have the budget to cover this expense" P25 |  | 3.4 What financial resources/budgets are required for the HPV vaccination programme over time? | Financial resources required |

Six individuals participated in follow-up interviews (5 in Mozambique; 1 based in North America or Europe). All of the participating individuals had remained in their roles since the first round of interviews.

### 4.1. Part 1: Evidence gap identification

Knowledge gaps were identified across four themes: (1) vaccine delivery strategy; (2) distributional impact of the vaccine programme; (3) vaccine economics; (4) comparison to other cervical cancer prevention methods. Table 1 presents interview extracts by theme (Part 1), mapped to research questions (Part 2). The third column presents outcome measures for each of the evidence needs. Numbers in parentheses in the following paragraphs refer to sub-theme numbers in Table 1.

#### 4.1.1. Vaccine delivery strategy

Based on multiple interviewee accounts, programme operationalization and the vaccine delivery strategy via schools were subject to a lengthy planning process after the HPV vaccine demonstration programme was concluded in 2015. The Ministry introduced the HPV vaccine in November 2021, when schools were closed, against the advice of the task force team in charge of programme planning. Due to global supply chain shortages of the vaccine, only a single age cohort (9-year-old girls) was targeted in the first year of vaccination. Study participants remained wary of the decision to introduce the vaccine in November 2021 and to focus on 9-year-old girls only. A study participant at a nationally-operating NGO was keen to explore the impact of vaccinating this single cohort only, (1.3):

> N, NGO: “Practically we’re not getting vaccines for the age group that we had initially intended [9 – 14-year-olds]. We’re just getting a single cohort. We’re going to do nine-year-old girls. So these are the things that we need to think about. And I think this is where the technical assistance needs to bring in these kinds of good practices into the ministry.” (P7)

Further, two study participants, one at the provincial level in government, and the other at locally-based international organization were interested in knowing the potential impact on population health if boys were also included in the target group (1.1). Study participants highlighted the need to evaluate the implications of targeting an older, sexually active age group, which included boys (1.2).

Beyond this, study participants were interested in how the HPV vaccine interacts with other primary prevention interventions, such as safer sex practices, as one study participant at an NGO (N) and a researcher at an international research institute highlighted (1.4).

#### 4.1.2. Distributional impact of the vaccine programme

Concerns around identifying and overcoming inequities in uptake were raised by several respondents. Interviewees at IOs flagged that the vaccination programme may not be accessible to hard-to-reach population groups, such as girls not attending schools or far away from healthcare facilities (2.1). For example, a study participant at an IO (HQ) highlighted that access to the vaccine is likely to be limited for girls, who do not attend state-run schools:

> HQ, IO: “The programme is really school-based, and it’s not very clear whether a very active attempt is made to deliver out of school. So, you know, some girls, including those in private schools, may just miss out on the vaccine because they’re not part of the public programme. […] So, I think, who we are reaching remains a black box. For example, nomadic populations in some parts of Africa are not reached, so, research is needed in that direction.” (P4)

Another study participant at an IO (HQ) voiced that other socioeconomic inequity indicators may be useful to inform a distributional impact analysis of the vaccine programme (P6). The concern of having varying vaccine coverage by rural and urban population sub-groups was also voiced by a study participant at a nationally-operating IO (P12). However, according to the study participant, this information could not be elicited from the vaccine coverage data.

#### 4.1.3. Vaccine economics

Study participants at national and international research institutes were interested in the cost, cost-effectiveness and financing of the vaccine (3.1). The cost per fully-immunized girl had previously been quantified by Alonso et al. ^48^ and Soi et al. ^40^. These cost estimates were derived from the demonstration programme experience in Mozambique. A study participant at a national research institute (P8) was, at the time of data collection, working on a new cost-effectiveness study, which relied on mathematical modelling. This mathematical model aimed to evaluate the cost-effectiveness of three HPV vaccines (CECOLIN, CERVARIX, and GARDASIL-4) in the medium and long term.^9^

The cost of transitioning to full domestic funding was raised by both global and national study participants. Specifically, interviewees at an IO (HQ, N) and the Ministry (N) wanted to understand how much the vaccine will cost once Gavi no longer supports the immunisation programme in Mozambique (3.2). The study participant at the Ministry voiced concerns about the sustainability of the vaccination programme:

> N, M: “There is the aspect of the price of vaccines for us as managers and so we are taking into account that until now almost all the vaccines that we are using in our system are financed by partners and there always comes that fear of: what will happen if one day the partners do not want to pay for these vaccines.” (P10)

Another study participant, at a nationally-operating NGO, flagged that vaccination programme costs should also be forecasted so that budgets could account for various aspects of the programme, e.g., equipment purchases (3.4).

Moreover, study participants at IOs (HQ), the Mozambican NITAG, and national research institutions were interested in learning about the vaccine’s wider benefit in Mozambique (3.3). For example, one study participant was interested in understanding the impact of the vaccine on the health sector, e.g., in terms of resources that will be saved due to fewer cervical cancer patients:

> N, IO: “If you reduce the number of women with cancer, you are also reducing the economic burden and investment in the area of treatment, so this is important to demonstrate.” (P13)

#### 4.1.4. Comparison to other cervical cancer prevention methods

Study participants were interested in how the HPV vaccine interacts with other primary prevention interventions, such as safer sex practices, as one study participant at an NGO and a researcher at an international research institute highlighted (4.1).

### 4.2. Part 2: Formulation of research questions

The evidence gap themes identified in interviews were translated into specific research questions, summarized in Table 1. Of these, five were selected for further exploration through modelling and evidence synthesis: the implications of a single-cohort strategy (1.3), the distributional impact of the vaccine programme (2.1), financing needs following the transition from Gavi support (3.2), the broader economic benefits of vaccination (3.3), and projected programme costs over time (3.4). To inform the feasibility and approach for addressing these questions, we reviewed the existing literature to examine how mathematical models have previously been applied to similar policy questions (Table 2).

**Table 2.** Examples of tools and mathematical modelling approaches to address knowledge gaps.

| **Questions posed** | **Examples of tools and mathematical modelling approaches applied in the peer-reviewed literature** |
| --- | --- |
| **1.2 What impact does sexual debut prior to vaccination have on the population health benefit (e.g., if older age groups are included in the target group)? How should the target group be defined with respect to sexual intercourse?** | HPV-specific, regional: Transmission- dynamic model with which the optimal age cohorts to vaccinate were identified.<sup>79</sup> |
| **2. 1 How do inequalities in access to education and infrastructure reflect in the vaccination coverage?** | HPV-specific, national: Economic evaluation of routine HPV vaccination quantifying distributional programme benefits following the Extended Cost Effectiveness Approach.<sup>80</sup> |
| **3.1 What is the cost-effectiveness of the different vaccine candidates?** | HPV-specific, global: Static models, dynamic economic models and a Markov model have been used to evaluate the vaccine on its cost-effectiveness in various contexts.<sup>81</sup> |
| **3.2 What costs does the vaccination programme imply following the transition from Gavi funding?** | A variety of vaccine programme costing studies are stored in a user-friendly database: https://immunizationeconomics.org/thinkwell-idcc |
| **3.3 What is the wider economic benefit of introducing the HPV vaccine?** | The definition of “wider economic benefit” needs to be determined together with the intended evidence user prior to identifying an appropriate modelling approach.<sup>11</sup> |
| **4.1 How does the vaccine introduction relate to/interact with/compare with other public health interventions and safe sex practices (e.g. condom use)?** | Not HPV-specific, global: A general model is presented to compute the effect of multiple, overlapping, and targeted interventions on epidemiological incidences.<sup>82</sup> |

### 4.3. Part 3: Evidence development

To address selected research questions, we applied and extended a closed cohort model to quantify the potential population-level impact of Mozambique’s HPV vaccination programme. The analysis focused on direct health benefits (in averted cervical cancer [CC] cases and deaths), financial protection for patients (through reduced out-of-pocket medical costs), health system efficiency (measured in full-time equivalents [FTEs] saved), and the equity of benefit distribution under both observed and hypothetical coverage scenarios.

At the observed 18.9% coverage level in 2021, the vaccination programme was estimated to avert up to 9.7% of expected CC cases and deaths, ranging from 4.0% in the highest socioeconomic quintile (HQ) to 11.6% in the lowest (LQ). Over the lifetime of a single vaccination cohort, this translates into an average of US$106,532 (range: US$48,382–$157,811) in treatment cost savings and the time equivalent of 59 full-time health workers saved annually. If 50% coverage were achieved and sustained over 30 years, an estimated 23.9% of CC cases and deaths could be prevented (LQ: 25.2%; HQ: 14.8%). Notably, in scenarios with pro-urban coverage gradients, rural populations in the lowest quintile were projected to benefit six times more than their urban counterparts.

These findings support evidence-informed planning and prioritization in Mozambique’s HPV vaccination strategy and may inform broader global policy discussions. A summary of the results was presented to stakeholders in an evidence brief (Figure 3), and full methodological details are published elsewhere.^31^

**Figure 3.**
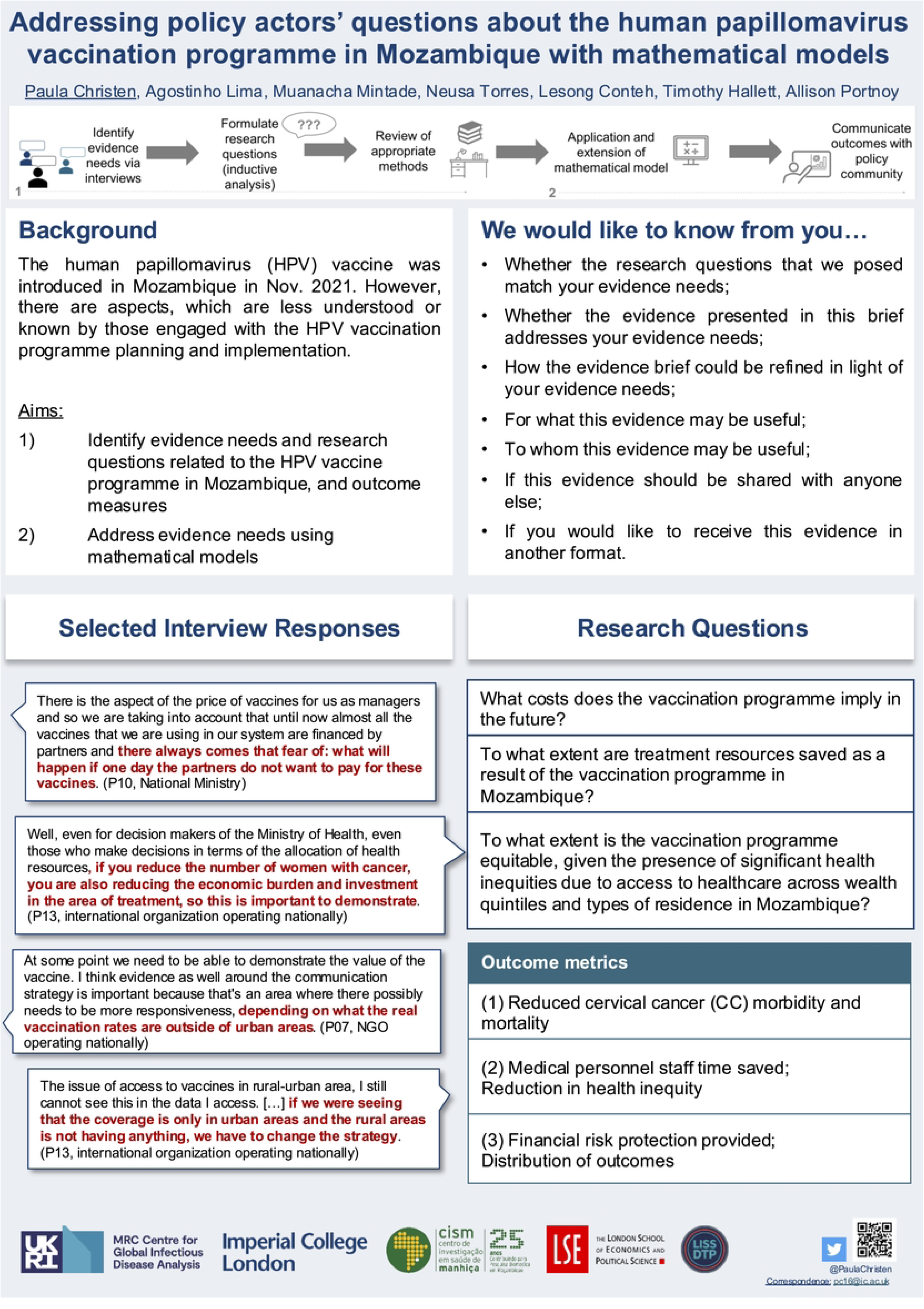
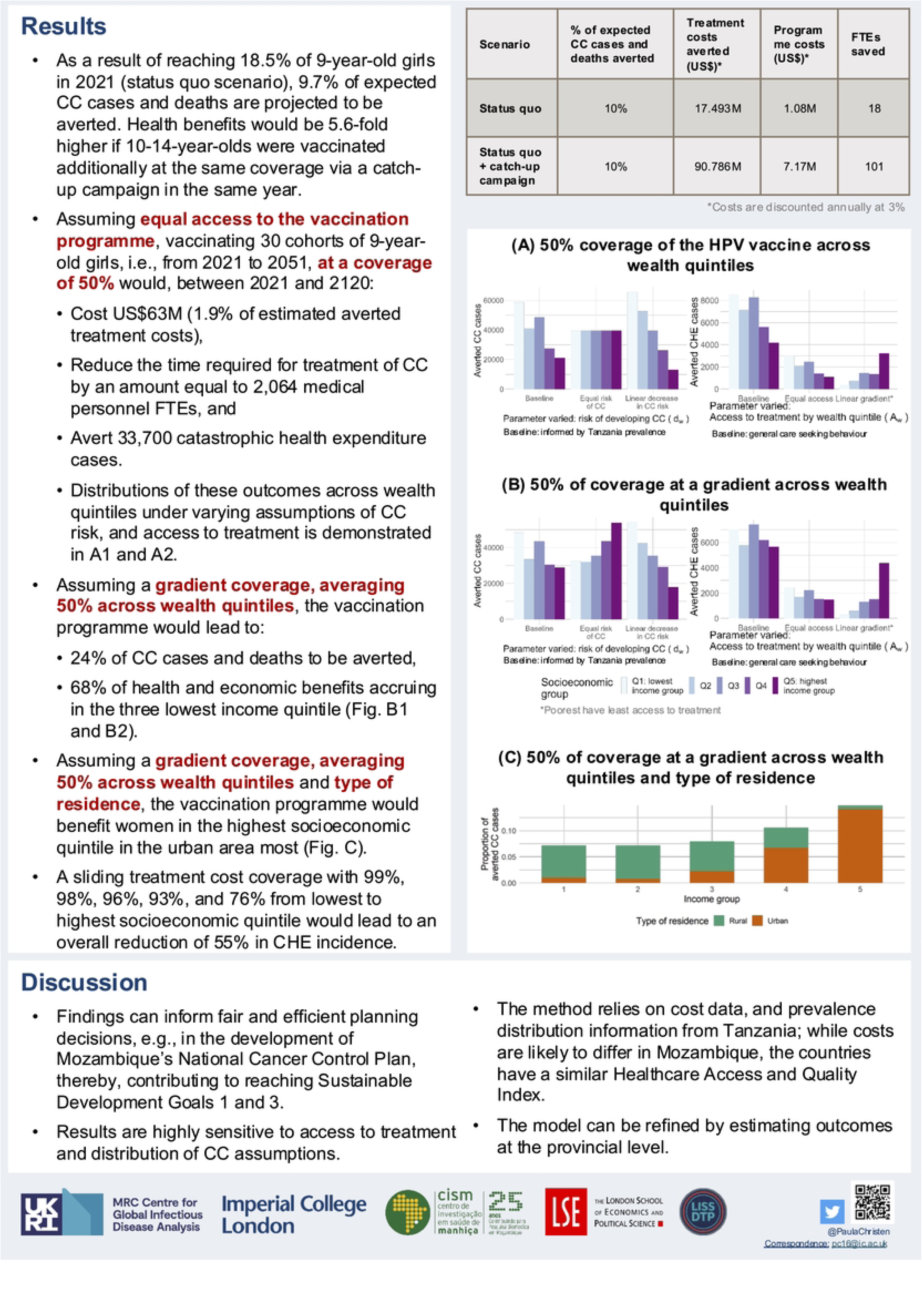
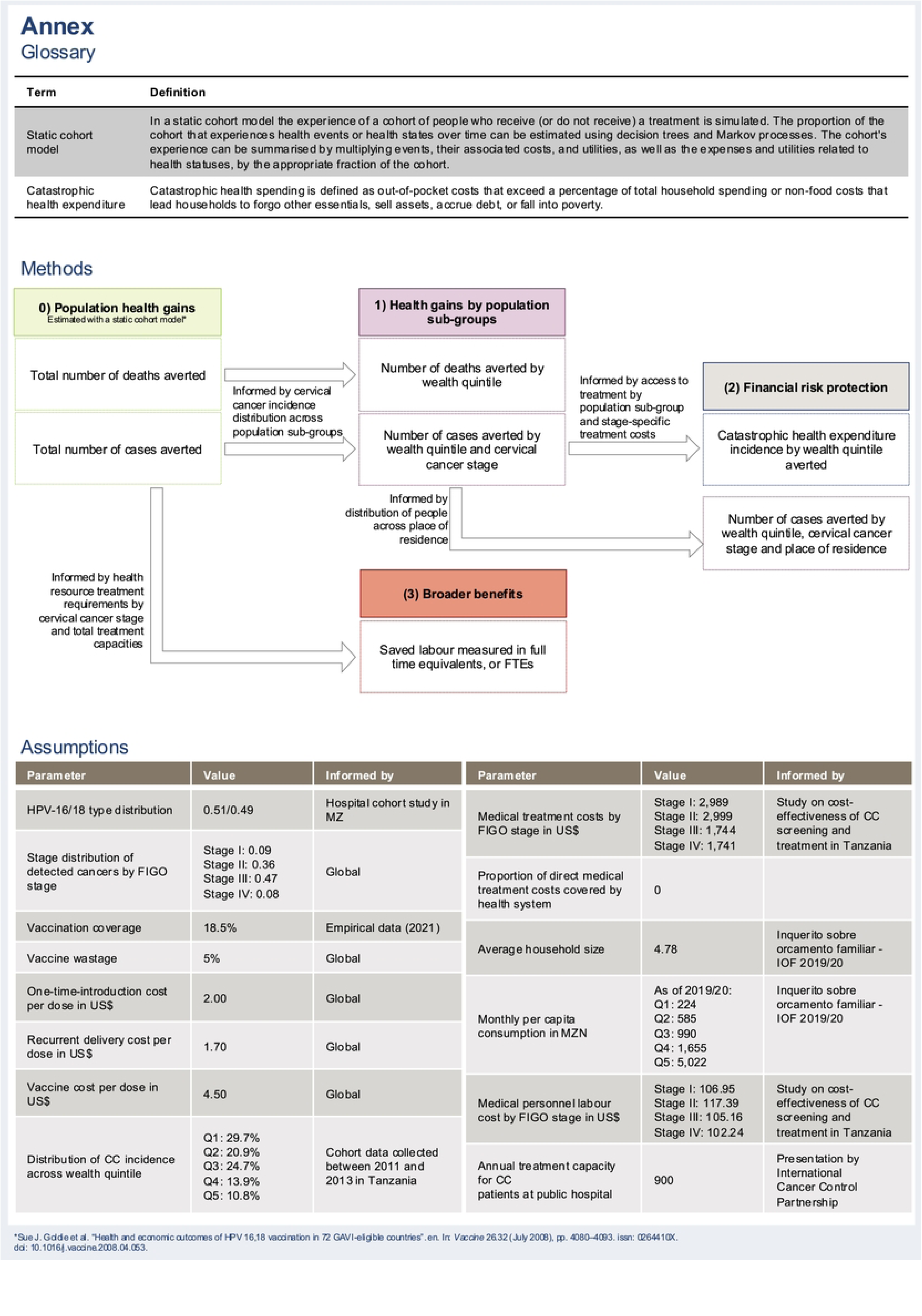
Evidence brief developed in Part 2 of the co-creation process.

This evidence brief summarizes stakeholder-identified priorities and corresponding research questions addressed using mathematical modelling. The brief was developed to communicate with policy actors engaged in HPV vaccination planning and implementation in Mozambique. Note: This figure comprises three parts spanning three pages.

### 4.4. Part 4: Reflections on utility of process and product

The final section delves into reflections on the evidence developed as presented in the evidence brief (Figure 3), its policy relevance, and utility. The following results are based on the views of the subset of eight study participants.

#### 4.4.1. Relevance of research questions and outcome measures

Study participants acknowledged the evidence presented in the evidence brief (Figure 3) to be relevant to their context. However, they did not recall having identified the questions addressed in the brief as reflecting their own knowledge gaps during the initial round of interviews. Although equity considerations—such as reaching underserved populations—emerged as a theme in earlier discussions, participants did not explicitly remember raising these issues. Nonetheless, they found the corresponding evidence highly relevant and valuable. They highlighted the importance of reaching girls in provinces with historically low coverage and of weighing the trade-offs between targeting vulnerable populations and the associated programme costs. To strengthen the analysis of equity in the context of the HPV vaccination programme, they suggested incorporating additional indicators (P6).

A key informant at the MOH emphasized the need to reduce the financial burden of HPV and cervical cancer on the healthcare system and families (P10). They discussed the aim of reducing morbidity and mortality from cervical cancer, as well as the potential cost savings associated with the vaccination intervention compared to the expenses of treating cervical cancer patients. The evidence brief is aligned with this concern in that it presents the vaccination programme costs and health sector costs foregone.

A key informant at the global level (P6) highlighted that other globally recognised tools (e.g., the WHO Prime tool^49^, the WHO Cervical Cancer Prevention and Control Costing (C4P) Tool^50^, and the HPV vaccine cost calculator^51^) could have been used to estimate vaccination programme costs. Thus, vaccination programme cost estimates were of less interest to them. Instead, the key informant voiced their interest in the other research questions addressed in the evidence brief.

All study participants criticized the coverage that served as the basis for the status-quo scenario as the one major premise. The coverage, which was informed by empirical data from the first three months of the vaccination programme, is not representative of the progress made. The study participants showed interest in receiving the estimates with updated coverage assumptions, as exemplified by the key informant:

> N, IO: “the only variable to update would be the coverage. We would have more realistic conclusions.” (P12)

All study participants confirmed that using data from neighbouring countries (e.g., Tanzania) was appropriate for informing parameter values used in the mathematical model. Participants did not have knowledge of other, possibly more appropriate, national sources for data gaps. One participant (P10) suggested speaking to colleagues at the MOH to explore national data sources that are not publicly available.

#### 4.4.2. Utility of evidence: Co-Identifying Policy-Relevant Research Questions

There was consensus that the vaccine impact estimates can inform resource allocation decisions, be used for financial planning, and support advocacy work in Mozambique. Interviewees felt that the distributional outcome analysis was particularly insightful and useful, especially in relation to strategies aiming to reduce the number of zero-dose children and missed communities, i.e., those that lack access to or are never reached by routine immunization services. As study participants learnt about how the evidence was developed and the outcomes were estimated, they voiced interest in refining the research, especially in its geographic level of analysis. When probing the utility of the findings, participants mostly commented on the utility of the evidence if it was presented at the provincial level but following the same analytical framework.

#### 4.4.3. Utility of evidence: Findings

##### Resource allocation

Analyzing the impact estimates across different socio-economic quintiles or regions can guide decision-makers on how to tailor strategies to specific situations. The modelling findings (as presented in the evidence brief) can support regional resource allocation decisions in identifying relevant outreach strategies that target specific socio-economic population sub-groups, as emphasized by a key informant at a nationally operating NGO:

> N, NGO: “Looking at the different quintiles we might be able to know for sure how to tackle a situation in specific region? Whether to involve more community health workers? How to best provide the vaccine, should it be via a health facility for people to come and get vaccinated? Should we go out to the communities?” (P23)

This was echoed by two key informants at a global organization and the MOH, who emphasized that the evidence quantify and highlight the issue for which tactics can then be developed in micro-planning activities, in which various factors such as out-of-school girls, vulnerable populations, immunocompromised individuals, and socioeconomic distributions are considered (P6, P10). The key informant at the MOH highlighted that this is particularly useful now as coverage rates are ‘inadequate’ and new strategies are under development. Strategies should consider the circumstances of and disparities across population sub-groups:

> N, IO: “In addressing equity issues in the programme we have identified the issue of hard-to-reach populations. The cyclone has affected different people in Mozambique differently, so there’s need to tailor strategies around that.” (P12)

This resonates with research question 2.1. While the evidence as presented in the evidence brief was seen as too high-level to inform programmatic decisions, the framework and outcome measures were considered policy-relevant.

Similarly, a key informant at a nationally-acting IO pointed out these findings are very relevant for “strategy preparation and the elaboration of campaigns” (P25). According to the participant, in this context, these findings can influence decision-makers to take a more holistic approach, considering various relevant aspects instead of relying solely on a global checklist.

##### Financial planning

According to key informant P23 (N, NGO), financial planning can also benefit from vaccine impact estimates. By providing detailed information on the cost of implementing different programmes, decision-makers can have a clearer understanding of the required financial resources. This level of detail would enable better budgeting decisions and the identification of areas where adjustments can be made to optimize resource allocation. Accurate cost estimates facilitate informed discussions on what to cut or adjust in the budgets allocated for immunization programmes. Examining the cost of vaccination per beneficiary, factors influencing these costs, and strategies to reduce them may also lead to less reliance on external funding:

N, NGO: “thereby, we might be able to reduce donor dependency so that the government can focus on other areas.” (P23)

##### Advocacy for resources

A key informant at the global level (P6) (HQ, IO) found the vaccine impact estimates served as a means to advocate for reaching under-immunized populations and promote equity in HPV vaccination. While the impact estimates demonstrate potential cost savings and the benefits of an equitable HPV vaccination programme, decision-makers need to consider the availability of human resources and domestic resources for outreach efforts. Therefore, increased domestic resources may ensure the sustainability of HPV programmes, acknowledging that Mozambique has made progress with over 50% coverage but still requires ongoing support to address the complexities of the vaccination programme.

##### Generalizability

Key informants at NGOs that also work with other countries felt that this type of analysis is also useful for programme planning in other countries that experience low vaccine coverage (P6).

##### Audience for evidence

Interviewees named various stakeholders who would benefit from receiving the vaccine impact estimates, including implementing partners, technical groups, MOH, and funding organizations. Important to note is that each stakeholder is interested in different aspects of the evidence presented. Interviewees recommended having selective meetings with specific individuals, such as the head of the immunization programme, the director of research, monitoring, and evaluation, and technical experts at the MOH, CoPI, and the Department for Planning and Corporation.

Further, programme managers, health system strengthening teams, and departments responsible for planning and cooperation were identified as relevant recipients of the vaccine impact estimates. This work may complement their efforts to planning activities targeting specific communities (P23) (N, NGO). One participant at a nationally-operating IO conveyed their enthusiasm about this work by searching for funding for a workshop on this research. This workshop would be held nationally and should engage all technical partner organizations.

At the international level, it was suggested to present this evidence to forums such as the Interagency Coordinating Committee, which is chaired by organizations including WHO, UNICEF, and other technical partners involved in immunization programmes (P13) (N, IO) and funding organizations, such as Gavi and the World Bank (P12) (N, IO).

## 5. Discussion

This paper set out to explore how collaborative engagement with evidence consumers can support the co-identification of policy-relevant research questions and the development of mathematical models to inform the HPV vaccine introduction in Mozambique. The findings highlight that identifying evidence gaps through dialogue with stakeholders can enhance both the relevance of the modelling process and its contribution to policy deliberation. Stakeholders’ reflections suggest that the joint framing of research questions and outcome measures helped ensure the evidence aligned with national priorities. In this case, population health, health sector, and societal perspectives were all considered relevant, even after the demonstration programme and lengthy programme planning. We find that through continued engagement beyond evidence development and by talking about the evidence, end-users could identify the utility of evidence and, together with the modellers, learn how it can inform policy decisions.

Unlike many modelling exercises that are agnostic of specific end-user needs, this method is uniquely tailored to the Mozambique setting by actively involving local stakeholders in defining both the model inputs and the desired policy-relevant outputs, ensuring that the evidence generated is directly applicable to policy dialogues. This type of co-production—grounded in shared problem-framing and iterative feedback—can maximize the policy relevance of mathematical modelling.^23,52,53^

Previous studies have furthered our understanding of how participatory approaches and stakeholder engagement can be employed in eliciting and depicting factors that should be accounted for in modelling exercises.^54,55^ Further, the literature advocates for platforms facilitating exchange and institutionalized multi-stakeholder approaches.^18,56^ To our knowledge, this is the first study to explore the use of interviews to identify knowledge gaps among policy and programme decision-makers in the context of epidemiological modelling to inform immunization programmes.

Integrating the perspectives of evidence consumers in the evidence development process led to evaluating the HPV vaccination programme from multiple perspectives, going beyond traditional health outcome measures (e.g., cases, deaths, or disability-adjusted life years averted). This study provides empirical evidence that aligns with conceptual frameworks emphasizing the importance of assuming a perspective in quantitative vaccine evaluations that consider the needs of decision-makers.^10^ The “Value of Vaccination” framework suggests considering both the payer perspective (e.g., health gains for the vaccinated) and the societal perspective (e.g., indirect health/economic gains).^57^ Other broader societal concepts, such as financial risk protection (FRP), peace of mind, societal health gains, political stability, social equity, and macroeconomic gains, are also gaining recognition as relevant vaccine benefits.^58^ Thus, frameworks such as the “Value of Vaccination” provide valuable guidance in the endeavour of developing policy relevant evidence.

Stakeholders also shared data not publicly available, illustrating how sustained engagement can uncover otherwise inaccessible resources.^59^ While not necessarily better, access to this data may potentially improve model utility and save time that might otherwise be spent identifying public data or establishing data-sharing agreements. There is an inherent trade-off between providing timely evidence and achieving perfect information.^60^ Investing time in dialogue with evidence consumers can offer guidance when navigating trade-offs — for example, between improving model accuracy and addressing newly emerging policy priorities. Over time, this process can generate a reinforcing cycle: stakeholder engagement yields more granular data, which enables more nuanced and policy-relevant findings, in turn increasing appetite for and investment in evidence use. This growing receptivity can help institutionalize more responsive and targeted analytic support in future decision-making.

Through such engagement, evidence producers and consumers are sensitized to each other’s environments, increasing the likelihood that evidence is used in policymaking.^61^ As a result, the issue of helicopter research^62^ can be mitigated. Echoing findings of previous research^15^, a bi-directional supply-demand relationship or dynamic is created through which follow-up questions about the evidence as well as new evidence needs may emerge. In line with previous research, frequent engagement between evidence producers and consumers can lead to trust in the evidence from mathematical models, confidence in using it to inform decisions, and ownership of the development and use process.^59,63^

Formally, engagement approaches such as the co-identification of research agendas may be best orchestrated by institutions that have been embedded in decision-making processes for a long period of time, such as the Expanded Programme for Immunization, the Manhiça Health Research Center (CISM), UNICEF Mozambique, and John Snow Inc in Mozambique. They have most knowledge of local decision-making processes, have oversight of stakeholders involved, and would therefore be best placed to convene a multi-stakeholder engagements.^64^

To support the co-identification of research questions and policy-relevant outcomes, quantitative skills to develop, adopt, and/or extend mathematical models of infectious diseases must be complemented with qualitative research skills to identify research questions and outcome measures. With the recent publication of an application of the PRIME model in Mozambique^65^, it appears that Universidade Eduardo Mondlane is developing technical epidemiological modelling capacity in Mozambique. Yet, ideally, evidence from multiple modelling groups would be considered to inform policy options and a multi-model comparison should be conducted prior to drawing policy recommendations.^66^

Eliciting research questions and outcome measures, objectively synthesizing recommendations stemming from research to inform policy options and facilitating the dialogue between evidence producers and consumers could be done by “knowledge brokers”.^67–69^ They would have to be skilled in quantitative and qualitative methods and have an understanding of decision-making processes in organizations that are engaged in vaccination programmes.^70^ Ideally, they can leverage formal organizational relationships to benefit from existing data-sharing agreements. They would not have to be a third-party service provider to mediate between organizations. Instead, organizations could identify champions in their organizations that can assume the role of knowledge brokers in their existing roles as evidence producers or consumers. Through secondments (evidence producers hosted by organizations that consume evidence and evidence consumers hosted by organizations that produce evidence, e.g., research institutions), the development and use of evidence from mathematical models, and more broadly advanced analytics, can be institutionalized, and capacities strengthened. Notably, in the context of vaccine introductions, stakeholders have vested political interests, and many lack impartiality.^71^ Thus, it is important to ensure that individuals do not have a conflict of interest when assuming the knowledge broker role.

In addition to institutional arrangements, interdisciplinary skill sets and capacity for implementation, effective co-identification and use of modelling evidence require sustained funding, multiple levels of engagement as well as negotiation.^21^ There appears to have been a movement among funders providing dedicated funding for participatory approaches ^21^, but more guidance should be provided on how they can be integrated effectively. The Mathematical and Economic Modelling for Vaccination and Immunisation Evaluation (MEMVIE) framework may be considered as one useful tool in guiding the implementation of stakeholder engagement in the context of developing evidence to inform vaccination policy recommendations specifically.^72^ The Co-creation Impact Compass is a more general framework that can be used for guidance in other healthcare research contexts as well.^73^

While this study focused on developing policy-relevant modelling evidence, future research should examine whether and how this research may have informed decisions in Mozambique. While complex to trace specific policy and programme decisions to a single piece of evidence, citations in grey literature as quantified by Overton, and qualitative accounts of knowledge translation as proposed by Kok et al. help to qualify knowledge translation.

Although the format of the evidence brief used in this study was informed by previous research identifying it as particularly helpful to policymakers for interpreting complex findings^74^, preferences likely vary depending on role, programme context, digital literacy, resource constraints, and the nature of the decision at hand. Exploring how to tailor evidence products to diverse user needs remains an important area for future work.

Moreover, it is crucial to keep in mind that, despite the encouragement to package policy recommendations so that consumers can easily understand and digest science, scientists must also remain objective and credible. Non-epistemic value judgements can compromise the epistemic integrity of scientific knowledge.^75^ At the same time, the ideal of value-free science and policy recommendations is “neither possible nor recommendable”^76^ as value-based decisions need to be made throughout the production of evidence, from the decision of what evidence need is addressed to the communication of findings.^77^ Instead, we propose that this process be made transparent so that epistemic and non-epistemic value-laden decisions can be subject to dialogue with evidence consumers. A stakeholder engagement approach, as explored in this research, enables mitigation of value-laden decisions throughout the research process.

### 5.1. Limitations

There are several limitations to this study. Firstly, interviews conducted in Portuguese had to be translated. As with all studies that use this process, perspectives can be misinterpreted or distorted. To mitigate this risk, the translator, MM, familiarized herself with topic-specific concepts in the areas of new vaccine introductions and evidence-informed decision-making. A subset of transcripts was back-translated into Portuguese to verify that the original meaning had been preserved. Further, all interviews were debriefed in regular team meetings such that interviewees could be contacted to answer follow-up questions if further clarification on their perspectives was required.

Secondly, stakeholder engagement in evidence development requires a high level of trust from the study participants as they are asked to share what they were uncertain or not knowledgeable about in their line of work. The highly trained and experienced interviewers tried to obtain the participants’ trust by reassuring them that their identity would be kept anonymous, and that information collected during interviews could always be revoked. Every participant had the opportunity to amend their answers post-interview, in case they felt they had shared confidential information, for example. When engaging stakeholders to developing evidence, it is advised to give participants enough time to think about their knowledge gaps, e.g., by sharing prompts ahead of data collection.

Thirdly, the interviews were time-constrained. This meant that study participants may have felt time-pressured, when trying to think what their priority evidence gaps were. Thus, participants were encouraged to reach out to the study team in case further questions would arise following the interview.

Fourthly, study participants were interested in more refined analyses. Further iterations of the process would lead to increased value in evidence and its utility. Ideally, the development of evidence would not end at this point but be continued in light of the participants’ feedback. As it stands, the model application only went through one iteration of gathering stakeholders’ input on research questions. The time points between interviews and follow-up interviews were >1 year, which meant that participants could not remember their knowledge gaps and did not directly identify with the quotes presented in the evidence brief. Moreover, the evidence needs may have changed since the first round of interviews, as the vaccination programme had already matured. Ideally, engagement is more frequent, e.g., through secondments of evidence producers/consumers and integration of knowledge brokers.

Moreover, before acting on evidence, it needs to be fully understood in terms of what it shows, what it does not show, and why, prior to decision-making. Effectively, evidence consumers need to have confidence in using evidence and making decisions with this information. The systematic evaluation of participants’ understanding of evidence was outside the scope of this research.

Study participants’ responses may have been subject to social desirability. Evidence consumers may have voiced their idealistic evidence use rather than the actual evidence use. We acknowledge that, beyond time constraints, there are multiple factors that can prevent evidence — including evidence based on more inclusive approaches like ours — from informing policy. However, investigating these barriers was beyond the scope of this study.^78^

Additionally, while the format of the evidence brief was designed to align with prior experiences and feedback, future research could explore which presentation formats are most accessible and actionable for different types of decision-makers. These preferences may vary by role, health programme, digital literacy, resource availability, and decision-making context.

## 6. Conclusion

In conclusion, this research has revealed that a stakeholder engagement in the development of mathematical modelling evidence for the HPV vaccination programme in Mozambique has substantial potential to enhance both the evidence development process and its utilization. By engaging stakeholders throughout the research process, the evidence generated was perceived by those involved in both policy development and implementation as more aligned to current in-country practical concerns and policy needs. This method led to the evaluation of the vaccination programme from multiple angles. By combining modelling with the insights of stakeholders directly involved in the HPV process in Mozambique, model inputs and outputs extended beyond the conventional health outcome measures typically used in modelling, offering a more contextually grounded understanding. This research underscores the importance of continuous dialogue between evidence producers and consumers, ensuring that the evidence is not only relevant but also trusted and owned by those who will use it to inform policy decisions.

## Data Availability

The data collected for the purpose of this study cannot be shared due to ethical restrictions.

## 7. Acknowledgements

The authors are deeply grateful to all study participants who generously contributed their time and insights, particularly amidst the challenges and competing demands of the COVID-19 pandemic. PC and TH acknowledge funding from the MRC Centre for Global Infectious Disease Analysis (reference MR/X020258/1), funded by the UK Medical Research Council (MRC). This UK funded award is carried out in the frame of the Global Health EDCTP3 Joint Undertaking. The study was part of a doctoral study programme (for PC) that was supported by the London Interdisciplinary Social Science Doctoral Training Programme (LISS DTP), funded by the Economic and Social Research Council (ESRC). Additional seed funding was provided by the MRC Centre for Global Infectious Disease Analysis to finance data collection in the country. The funding sources had no involvement in conducting the research.

## 8. Declaration of generative AI and AI-assisted technologies in the writing process

During the preparation of this work the authors used ChatGPT 4.o in order to improve text clarity. After using this tool/service, the authors reviewed and edited the content as needed and take full responsibility for the content of the publication.

